# Clinical Pharmacy Services Provided in Public Sector Hospitals in Nigeria: A National Survey

**DOI:** 10.1101/2021.06.17.21258942

**Authors:** Arit Udoh, Mary Akpan, Umar Idris Ibrahim, Basira Kankia Lawal, Kamilu Sarki Labaran, Ekpedeme Ndem, Kosisochi Amorha, Ayodeji Matuluko, Olubukola Tikare, Unoma Ohabunwa, Eneyi Kpokiri

## Abstract

**Background:** Studies show that clinical pharmacy services are effective in optimizing medicines use and patients’ outcomes. This study aimed to determine the clinical pharmacy services provided in public sector hospitals in Nigeria.

**Methods:** This was an online survey of 296 primary, secondary and tertiary care hospitals sampled purposively across the 36 States and Federal Capital Territory in Nigeria. Data analysis was conducted descriptively, and via Chi-square test and multivariate analysis of variance (MANOVA).

**Key findings:** Responses were obtained from 272 hospitals in the country with a survey completion rate of 88%. This included 55 tertiary, 72 secondary, and 145 primary healthcare centres (PHCs). Pharmacists provided pharmaceutical care services in all the tertiary care hospitals, 94% of the secondary, and in only 6% of the PHCs surveyed. The composition of the pharmacy department per level of care was similar across the six geopolitical regions (V = 0.383, F = 1.453, P = 0.06) with more pharmacy staff employed in the tertiary care hospitals compared to the secondary care facilities. The majority (≥75%) of the tertiary and secondary care hospitals provided medicines information, patient education and counselling, alongside in- and outpatient dispensing services. However, fewer than 30% reported routine pharmacists’ involvement in multidisciplinary ward rounds, medication chart review, therapeutic guidelines development, antibiotic stewardship programmes, and drug therapy monitoring. Pharmacists routinely provided medication error reporting services in only about half of the hospitals in the sample, and this was not associated with the level of care (p > 0.05).

**Conclusions:** The findings of this study demonstrate disparity in clinical pharmacy service availability across the various levels of care in Nigeria. It also highlights the need to scale up and prioritize their integration within the primary care sector.

## INTRODUCTION

In November 2019, the National Council on Establishments (NCE) approved the consultant pharmacist cadre for inclusion in the public sector schemes of service in Nigeria [1]. This approval was assented to by the Head of the Federal Civil Service in September 2020 [1], and represents government acknowledgement of the essential role of pharmacists as medicines experts within the multidisciplinary clinical team. This acknowledgement underscores pharmacists’ evolving roles from the focus on medicines and medicinal products, to the provision of patient-oriented services. Globally, this patient-oriented practice model, which is the concept of clinical pharmacy; has been shown to optimize medicines use, assure medication safety, and improve patients’ outcomes and quality of life [2].

Although clinical pharmacy is well established in several high income countries [3–7]; its uptake has varied across the world regions and within nations [8–11]. Until recently, the delivery of these services in countries in Africa has been limited and/or non-existent [12–14]. The revised Basel Statements on the Future of Hospital Pharmacy, published in 2014, emphasized the pharmacists’ role in the clinical team, including their influence on prescribing and monitoring of medicines use [15,16]. The increasing involvement in patient-oriented service provision underscores the imperative for the availability of appropriately trained pharmacists equipped with the requisite skills needed to provide enhanced pharmaceutical care.

In Nigeria, estimates show that hospital pharmacists make up about 20% of the licensed pharmacy workforce [17]. Although data on the distribution of pharmacists across the private and public sector hospitals in the country is lacking in the literature; anecdotally, majority of the hospital pharmacists are employed in the public sector facilities. Also, most of the internship and post-graduate clinical training of pharmacists are carried out at these hospitals. However, information on the pharmaceutical care services provided, and the involvement of hospital pharmacists in patient-oriented care is unclear. A previous study of hospital pharmacy practice in four States in the North Western region of Nigeria estimates that pharmaceutical care services are provided in about 47 - 85% of the facilities surveyed per state [18]. This information is missing in the literature for the other parts of the country. Given the paucity of data, this study aimed to determine the clinical pharmacy services provided in public sector hospitals in Nigeria. The findings of this study will inform national policy planning in relation to these services.

## METHOD

### Study setting

Nigeria is the most populous country in Africa with a population of approximately 200 million [19]. The country is divided in to six geopolitical zones and comprise the Federal Capital Territory (FCT), 36 States, and 774 Local Governments Areas (LGAs). About two-thirds of the hospitals in the country are government owned with the public sector primary, secondary and tertiary care facilities funded by the local, state and federal governments, respectively [20,21]. Government owned primary care facilities are mainly situated in rural areas and comprise the health centres and health posts [22]. In contrast, public sector secondary and tertiary care facilities are predominantly located in urban/peri-urban areas. The district and general hospitals make up the secondary care facilities in the country while tertiary care providers include the teaching hospitals, federal medical centres (FMCs), and other federal funded facilities. The public sector tertiary care facilities are designed to serve as referral hospitals for complex and specialised care provision while the primary care facilities provide basic and preventative health care services [23,24]. However, poor funding, limited health infrastructure, as well as staffing and medicines shortages at the primary and in most secondary care hospitals, has led to a preference for tertiary care service providers in population health seeking in Nigeria [24].

### Study design

This study was an online survey of public sector hospitals in Nigeria. The survey included public sector primary, secondary and tertiary care hospitals sampled across the 36 States and the FCT. To ensure that all the states and FCT in the country were duly represented in the survey; the hospitals per state were sampled purposively. Specifically, the Nigeria Health Facility Registry of the Federal Ministry of Health [25] was consulted to identify the public sector hospitals in the country. Thereafter, all the teaching hospitals and FMCs listed were targeted for data collection. Where feasible, data from an additional federal government funded facility in each State were also collected to make up two tertiary care hospitals. In addition, two general hospitals and four primary healthcare centres (PHCs)/health posts in each of the states were targeted. In total, 74 tertiary and secondary care facilities each, and 148 PHCs/health posts were targeted.

### Data collection

Data collection was via the online Qualtrics® software (Qualtrics, Utah, USA). The hospital administrator, head of the pharmacy department or a designated senior pharmacist with managerial responsibility at each target facility provided data for the survey. Respondents for this survey were identified via the Association of Hospital and Administrative Pharmacists of Nigeria (AHAPN), the state chapters of the Pharmaceutical Society of Nigeria (PSN), and through the authors’ professional network. The online survey link was shared with each respondent with consent to participate required prior to data completion. This survey was conducted over 12 months and data collection concluded on 30 July 2020. Ethical review and approval for this study was obtained from the National Health Research Ethics Committee (NHREC) of the Federal Ministry of Health in Nigeria (REF NO: NHREC/01/01/2007).

### Survey Instrument

The data collection tool (provided in Appendix 1) was adapted from a previously validated instrument used in a survey of hospital pharmacy practice in Ireland [7]. The adapted questionnaire was pre-tested for face and content validity in a sample of five pharmacists who were not directly involved in this study. Feedback obtained from the pre-test was incorporated with further iteration resulting in a questionnaire comprising 23 items that required a combination of multiple choice, “Yes”, “No”, “Not applicable”, or free text responses. The survey questions were presented over five pages. The first page of the questionnaire included a consent question that required an affirmative answer to be eligible to participate.

To ensure completeness, all the survey questions were mandatory. Respondents had to provide an answer to the questions on a given page in order to proceed to the next. Given the objective of this study and to ensure a meaningful interpretation of the survey findings; a filtering question was employed. The provision of pharmaceutical care services by pharmacist at each facility was the filtering question used. This question was presented in the demographic section on page 2, and respondents who answered “No” were automatically taken to the end of the survey.

### Data Analysis

Quantitative data obtained in this study were analysed using SPSS v26 (IBM, USA). Descriptive statistics including frequency (counts, percentages), mean (standard deviation (SD)), median (interquartile range (IQR)) were used to summarize the data while the Pearson’s Chi-square (*X*^*2*^) was used to assess association between categorical variables. Disparity in the composition of the pharmacy department and the availability of clinical pharmacy services per level of care across the six geopolitical zones in the country, was assessed using the Pillai’s Trace statistic (*V*) of the multivariate analysis of variance (MANOVA). The Pillai’s Trace multivariate statistic was chosen because it is more robust to outliers and violation of normality [26]. Confirmatory post-hoc analysis was also conducted using the Bonferroni correction. The findings of this survey are reported in line with the Checklist for Reporting Results of Internet E-Surveys (CHERRIES) guidelines [27].

## RESULTS

In total, 308 respondents accessed the online survey link on Qualtrics and consented to participate in the study. Of this number, 272 provided complete responses to the survey questions, indicating a completion rate of 88%. The 36 incomplete responses included those who consented to participate on the first page of the questionnaire but did not attempt the survey questions (n=11), and others who attempted only some of the demographic questions (n=25). The incomplete responses were not useable and were therefore excluded from further analysis.

### Demography

Of the 272 complete responses obtained, 55 were from tertiary care facilities and comprised all the teaching hospitals, FMCs and 11 other government-funded hospitals in the country; 72 were secondary care providers and these were all general hospitals, while the remaining 145 were PHCs. Geographically, responses were obtained from hospitals across the 36 States in the country including the FCT. Regionally, the highest number of responses were from hospitals in the North West (n = 51, 19%) while the least were from the South East (n = 36, 13%) regions (Table 1).

**Table 1:** Hospital Profile and demography.

| Hospital Profile |  | Hospital Level of Care |  |  |
| --- | --- | --- | --- | --- |
|  |  | Primary<br>(N=145) | Secondary<br>(N=72) | Tertiary<br>(N=55) |
| Geographical distribution,<br>n (%) | North Central | 28 (19) | 11 (15) | 9 (16) |
|  | North East | 23 (16) | 12 (17) | 8 (15) |
|  | North West | 27 (19) | 14 (19) | 10 (18) |
|  | South East | 20 (14) | 9 (13) | 7 (13) |
|  | South South | 23 (16) | 12 (17) | 11 (20) |
|  | South West | 24 (16) | 14 (19) | 10 (18) |
| Size of facility<br>(Number of beds, n (%)) | <100 beds | 145 (100) | 54 (75) | 6 (11) |
|  | 101 – 250 | 0 (0) | 18 (25) | 16 (29) |
|  | 251 – 500 | 0 (0) | 0 (0) | 24 (44) |
|  | ≥ 501 | 0 (0) | 0 (0) | 9 (16) |
|  | Median N (IQR) | 15 (20) | 100 (77) | 368 (187) |
|  | Min – Max | 6 – 30 | 12 – 250 | 60 – 850 |
| Pharmacy opening times,<br>n (%) | < 24 hours | 145 (100) | 41 (57) | 10 (18) |
|  | 24 hours | 0 (0) | 31 (43) | 45 (82) |
| Number of dispensing<br>locations at facility, n (%) | 1 – 3 | 145 (100) | 52 (72) | 6 (11) |
|  | 4 – 6 | 0 (0) | 20 (28) | 15 (27) |
|  | > 6 | 0 (0) | 0 (0) | 34 (62) |
| Pharmacist provide<br>pharmaceutical care<br>services at facility, n (%) | Yes | 9 (6) | 68 (94) | 55 (100) |
|  | No | 136 (94) | 4 (6) | 0 (0) |

### Hospital profile

Median hospital size with respect to number of beds varied across the three levels of care. The tertiary care facilities in the sample were generally larger compared to the secondary and primary care hospitals (Table 1). The size of the hospital was also reflected by the number of dispensing locations available. The PHCs and about 72% of the secondary care facilities had between one to three dispensing locations for pharmaceuticals while the majority (89%) of the tertiary care facilities had about four or more (Table 1). Pharmaceutical care services were provided by pharmacists in all (100%) of the tertiary facilities, and in most of the secondary care hospitals (N = 68, 94%). However, pharmacists were only available in fewer than 10% of the PHCs in this survey (Table 1). Given the objective of this study, which was to determine the clinical pharmacy services provided; only the data obtained in the tertiary and secondary care facilities were further analysed hereafter.

### Pharmacy department profile

This section of the analysis included all the tertiary and the 68 secondary care hospitals that reported that pharmaceutical care services were provided by pharmacists (N = 123). Generally, the composition of the pharmacy department was comparative across the six geopolitical regions in the country (V = 0.383, F = 1.453, P = 0.06) with no statistical significant difference observed beyond the level of care provided in the respective hospitals (V = 0.522, F = 18.378, P = 0.001). The total number of pharmacy staff in the respective cohorts varied with more pharmacists, pharmacy technicians and support staff employed in the tertiary care hospitals compared to the secondary care facilities (Table 2). This was reflective of the size of the hospitals in the respective cohorts. More than 70% of the pharmacists employed in the tertiary care hospitals were those with five or more years of practice experience, compared to the secondary care cohort with about 57% (Table 2). A higher proportion of the pharmacists in the tertiary care cohort had a post-graduate degree or professional recognition compared to the secondary care with about a third (Table 2).

**Table 2:**
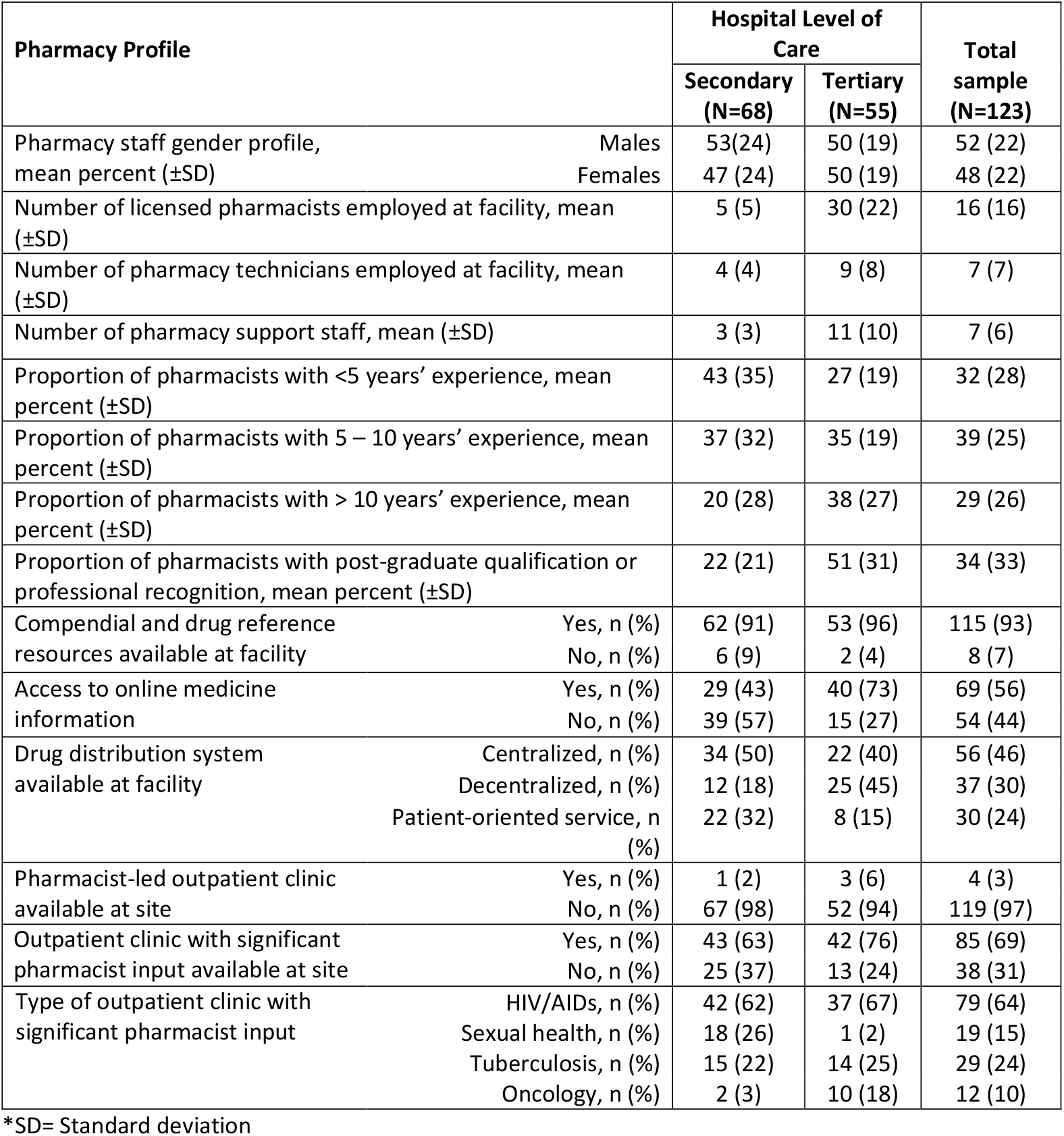
Pharmacy department profile per level of care.

| Pharmacy Profile |  | Hospital Level of Care |  | Total sample (N=123) |
| --- | --- | --- | --- | --- |
|  |  | Secondary (N=68) | Tertiary (N=55) |  |
| Pharmacy staff gender profile, mean percent (±SD) | Males | 53(24) | 50 (19) | 52 (22) |
|  | Females | 47 (24) | 50 (19) | 48 (22) |
| Number of licensed pharmacists employed at facility, mean (±SD) | | 5 (5) | 30 (22) | 16 (16) |
| Number of pharmacy technicians employed at facility, mean (±SD) | | 4 (4) | 9 (8) | 7 (7) |
| Number of pharmacy support staff, mean (±SD) | | 3 (3) | 11 (10) | 7 (6) |
| Proportion of pharmacists with <5 years' experience, mean percent (±SD) | | 43 (35) | 27 (19) | 32 (28) |
| Proportion of pharmacists with 5 – 10 years' experience, mean percent (±SD) | | 37 (32) | 35 (19) | 39 (25) |
| Proportion of pharmacists with > 10 years' experience, mean percent (±SD) | | 20 (28) | 38 (27) | 29 (26) |
| Proportion of pharmacists with post-graduate qualification or professional recognition, mean percent (±SD) | | 22 (21) | 51 (31) | 34 (33) |
| Compendial and drug reference resources available at facility | Yes, n (%) | 62 (91) | 53 (96) | 115 (93) |
|  | No, n (%) | 6 (9) | 2 (4) | 8 (7) |
| Access to online medicine information | Yes, n (%) | 29 (43) | 40 (73) | 69 (56) |
|  | No, n (%) | 39 (57) | 15 (27) | 54 (44) |
| Drug distribution system available at facility | Centralized, n (%) | 34 (50) | 22 (40) | 56 (46) |
|  | Decentralized, n (%) | 12 (18) | 25 (45) | 37 (30) |
|  | Patient-oriented service, n (%) | 22 (32) | 8 (15) | 30 (24) |
| Pharmacist-led outpatient clinic available at site | Yes, n (%) | 1 (2) | 3 (6) | 4 (3) |
|  | No, n (%) | 67 (98) | 52 (94) | 119 (97) |
| Outpatient clinic with significant pharmacist input available at site | Yes, n (%) | 43 (63) | 42 (76) | 85 (69) |
|  | No, n (%) | 25 (37) | 13 (24) | 38 (31) |
| Type of outpatient clinic with significant pharmacist input | HIV/AIDs, n (%) | 42 (62) | 37 (67) | 79 (64) |
|  | Sexual health, n (%) | 18 (26) | 1 (2) | 19 (15) |
|  | Tuberculosis, n (%) | 15 (22) | 14 (25) | 29 (24) |
|  | Oncology, n (%) | 2 (3) | 10 (18) | 12 (10) |

\*SD= Standard deviation

About half of the secondary care cohort reported a centralized drug distribution system compared to the approximately 60% in the tertiary care respondents that reported a decentralized or a patient-oriented drug distribution system (Table 2). Pharmacist-led outpatient clinics were available in only four (3%) hospitals in the sample. Relative to the secondary care hospitals, other outpatient clinics with significant pharmacists’ input beyond dispensing were reported in more of the tertiary care hospitals (Table 2), however, this was not statistically significant (X^2^ = 2.45, P = 0.12) (Table 2). On the other hand, sexual health clinics with significant pharmacists’ input were more likely to be available in secondary care (X^2^ = 13.59, P = 0.001), while oncology clinics were more likely to be reported in the tertiary care hospitals (X^2^ = 8.46, P = 0.004) (Table 2).

### Clinical pharmacy services provided

The study showed that medicines information, patient education and counselling, in-patient and outpatient dispensing services were always available in the majority (≥ 75%) of the tertiary and secondary care hospitals in the sample (Table 3). However, fewer than a third of the respondents in either cohort indicated that pharmacists were routinely involved in multidisciplinary ward rounds, therapeutic guidelines development, antibiotic stewardship programmes, anticoagulant services, drug therapy monitoring, clinical trials, and parenteral nutrition service provision (Table 3). Comparatively, discharge counselling, drug utilization evaluation and review, compounding/extemporaneous preparations, and medical device services were more likely to be available in the tertiary care hospitals while vaccines services were more likely to be provided in the secondary care hospitals (p<0.05) (Table 3). Only about half of the hospitals in the overall sample reported the availability of a medication error, pharmacovigilance/adverse drug events (ADE) or adverse drug reaction (ADR) reporting service, and this was not associated with the level of care (p > 0.05) (Table 3).

**Table 3:** Availability of clinical pharmacy services per level of care. (*statistical significance p < 0.05)

| Clinical pharmacy services provided | Hospital Level of Care |  |  |  |  |  |  |  |  |  |  |  | X<sup>2</sup> | P |
| --- | --- | --- | --- | --- | --- | --- | --- | --- | --- | --- | --- | --- | --- | --- |
|  | Secondary (N=68) |  |  |  |  |  | Tertiary (N=55) |  |  |  |  |  |  |  |
|  | Never |  | Rarely |  | Always |  | Never |  | Rarely |  | Always |  |  |  |
|  | n | % | n | % | n | % | n | % | n | % | n | % |  |  |
| Multidisciplinary ward rounds | 31 | 46 | 28 | 41 | 9 | 13 | 27 | 49 | 19 | 35 | 9 | 16 | 0.574 | 0.751 |
| Medication Chart review | 25 | 37 | 23 | 34 | 20 | 29 | 18 | 33 | 24 | 44 | 13 | 24 | 1.398 | 0.491 |
| Medication error reporting | 9 | 13 | 23 | 34 | 36 | 53 | 5 | 9 | 24 | 44 | 26 | 47 | 1.742 | 0.481 |
| Therapeutic guidelines and protocol development | 26 | 38 | 28 | 41 | 14 | 21 | 22 | 40 | 22 | 40 | 11 | 20 | 0.01 | 0.995 |
| Inpatient dispensing | 12 | 18 | 13 | 19 | 43 | 63 | 7 | 13 | 3 | 5 | 45 | 82 | 6.599 | 0.037* |
| Outpatient dispensing | 0 | 0 | 0 | 0 | 68 | 100 | 0 | 0 | 0 | 0 | 55 | 100 | 0 | 1 |
| Medicines information | 0 | 0 | 10 | 15 | 58 | 85 | 0 | 0 | 11 | 22 | 39 | 78 | 3.107 | 0.078 |
| Antimicrobial stewardship | 35 | 51 | 26 | 38 | 7 | 10 | 18 | 33 | 22 | 40 | 15 | 27 | 7.761 | 0.021* |
| Contraceptive services | 35 | 51 | 17 | 25 | 16 | 24 | 26 | 47 | 18 | 33 | 11 | 20 | 0.999 | 0.607 |
| Vaccines services | 13 | 19 | 12 | 18 | 43 | 63 | 19 | 35 | 19 | 35 | 17 | 31 | 12.179 | 0.002* |
| Cold chain management | 9 | 13 | 10 | 15 | 49 | 72 | 11 | 20 | 8 | 15 | 36 | 65 | 1.135 | 0.567 |
| Anticoagulant services | 46 | 68 | 16 | 24 | 6 | 9 | 25 | 45 | 14 | 25 | 16 | 29 | 9.947 | 0.007* |
| Medical devices services | 31 | 46 | 14 | 21 | 23 | 34 | 8 | 15 | 22 | 40 | 25 | 45 | 14.87 | 0.001* |
| Compounding/Extemporaneous preparation | 18 | 26 | 32 | 47 | 18 | 26 | 4 | 7 | 4 | 7 | 47 | 85 | 43.472 | 0.001* |
| Pharmacovigilance/ADE/ADR reporting | 5 | 7 | 33 | 49 | 30 | 44 | 2 | 4 | 23 | 42 | 30 | 55 | 1.851 | 0.396 |
| Drug therapy monitoring | 8 | 12 | 36 | 53 | 24 | 35 | 9 | 16 | 28 | 51 | 18 | 33 | 0.589 | 0.745 |
| Medicines reconciliation/history | 13 | 19 | 29 | 43 | 26 | 38 | 12 | 22 | 14 | 25 | 29 | 53 | 4.499 | 0.105 |
| Involvement in clinical trials | 54 | 79 | 14 | 21 | 0 | 0 | 23 | 42 | 22 | 40 | 10 | 18 | 23.143 | 0.001* |
| Patient education & counselling | 0 | 0 | 3 | 4 | 65 | 96 | 0 | 0 | 4 | 7 | 51 | 93 | 0.464 | 0.496 |
| Discharge counselling | 18 | 26 | 29 | 43 | 21 | 31 | 7 | 13 | 17 | 31 | 31 | 56 | 8.616 | 0.013* |
| Drug utilization evaluation & review | 20 | 29 | 29 | 43 | 19 | 28 | 6 | 11 | 19 | 35 | 30 | 55 | 10.838 | 0.004* |
| Aseptic services | 34 | 50 | 22 | 32 | 12 | 18 | 16 | 29 | 23 | 42 | 16 | 29 | 5.764 | 0.056 |
| Parenteral nutrition support (TPN and enteral feeds) | 41 | 60 | 15 | 22 | 12 | 18 | 23 | 42 | 18 | 33 | 14 | 25 | 4.162 | 0.125 |

The results also showed disparity in the availability of clinical pharmacy services across the six geopolitical regions (V = 1.293, F = 1.570, P = 0.001). Post-hoc analysis indicated that pharmacists in more of the North Central and South West hospitals (n = 5(26%), 9(36%), respectively) were routinely involved in multidisciplinary ward rounds compared to the other regions with fewer than 5% each. The North Central, South West and South South respondents were more likely to respectively report routine availability of medication chart review (n = 12(58%), 8(35%), 10(43%)) and therapeutic guideline development services (n = 6(32%), 6(26%), 8(35%)), compared to the other regions with fewer than 10% each. Although most of the hospitals in the North Central, North East, North West and South West regions provided compounding/extemporaneous preparation services (n = 16(79%), 11(56%), 12(52%), 12(52%), respectively); fewer than half of those in the South East and South South (n = 5(31%), 10(43%), respectively) reported this. Medication error services were reported by more than half of the North Central, North East, South South and South West (n = 13(63%), 11(55%), 12(52%), 17(70%), respectively) respondents compared with only about a third in the North West and South East region (n = 7(30%), 4(25%), respectively).

### Respondents’ perception of practice

Overall, only a few of the respondents (≤ 30%) agreed that the available technology in the pharmacy department, the continuous professional development (CPD) opportunities, and the pharmacists’ influence on prescribing was adequate or satisfactory at the respective hospitals (Table 4). This perception was not associated with the level of care provided in the hospitals (p > 0.05). More than half of those in the tertiary care compared to a smaller proportion in the secondary care hospitals, agreed that the pharmacy department had the right skills mix with satisfactory opportunities for developing expertise in hospital pharmacy practice (Table 4). However, this difference in perception was not statistically Significant (p > 0.05) (Table 4). The majority of the tertiary (n = 48(87%)) and secondary care (n =53 (78%)) respondents agreed that interdisciplinary collaboration in the hospital will raise the profile of the pharmacy department (Table 4). Generally, respondents’ perceptions of practice within the respective levels of care were comparative across the six geopolitical regions (V = 0.333, F = 0.888, p = 0.681).

**Table 4:**
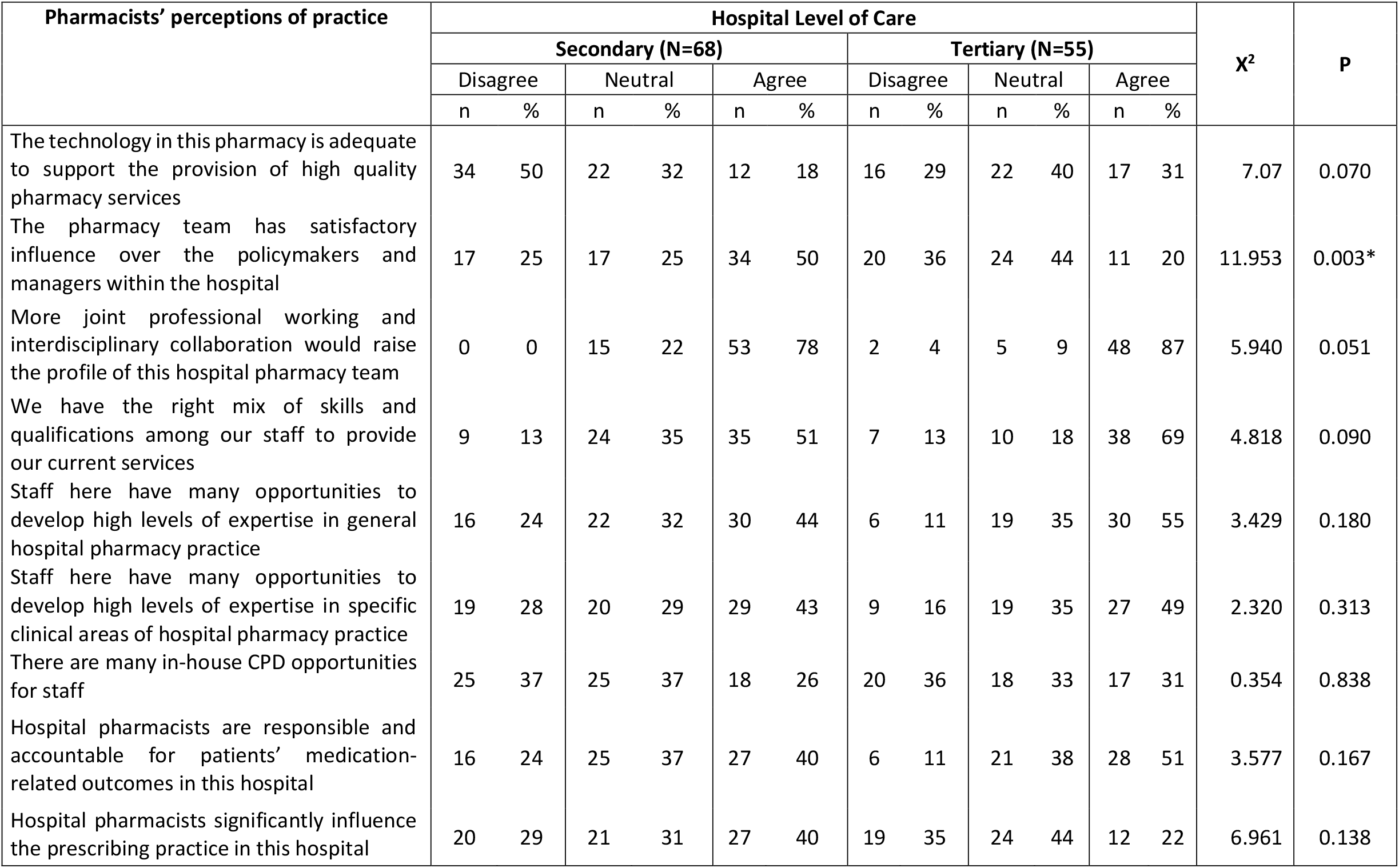
Respondents’ perceptions of practice and available resources. (*statistical significance p < 0.05)

## DISCUSSION

Pharmacists’ are essential for attaining the goal of universal health coverage and equitable access to essential health services, particularly with respect to optimizing the safe and responsible use of medicines [28]. Existing reports indicate that pharmaceutical care services and medicines-related activities are carried out by non-pharmacists in the majority of the PHCs in Nigeria [23,29,30]. This was observed in our study as pharmacists were employed in only a fraction of the primary care facilities. This finding highlights the need to prioritize the integration of clinical pharmacists within the primary care system in Nigeria. This is essential, given the evidence from other countries that demonstrate the effectiveness of pharmacist- led primary care interventions in long term disease prevention, medication therapy management, and improvement in drug-related patient outcomes [31,32].

Although outpatient clinics with significant pharmacist input beyond dispensing were available in about two-thirds of the hospitals in our survey; the disparity in service availability within the specific levels of care and across the various geopolitical regions indicate the need for national scale up. Several factors including significant workforce shortages, high attrition rate, poor remuneration and funding, and the available pharmacists’ expertise are some of the factors reported to limit uptake of clinical pharmacy services in countries in Africa including Nigeria [9,12,18,33–35]. This may explain why some of the clinical pharmacy services assessed in this survey were more likely to be available in the tertiary facilities compared to the secondary care hospitals; especially as the former tend to be larger, more funded and equipped with the capacity of employing more staff. On the other hand, this may also be related to the practice experience of the licensed pharmacists employed in the hospitals within the respective levels of care. Pharmacists in the tertiary care hospitals tended to be more experienced with a larger proportion possessing a post-graduate qualification or professional recognition. This suggests that the tertiary care pharmacists are more likely to have undertaken further post-registration training and are potentially more equipped to provide enhanced patient-oriented care services in their respective hospitals.

The finding that online medicine information services were not available in close to half of the secondary care hospitals highlights the need to improve the available information technology infrastructure in the respective hospitals. This is necessary, so as to ensure pharmacists’ access to current medicines information resources, especially as existing evidence demonstrates that professional practice that is consistent with up-to-date knowledge influence patient safety and clinical outcomes [36]. This was also emphasized by the significant proportion of study respondents in both the tertiary and secondary care hospitals who indicated that the IT resources and CPD opportunities available were not adequate. More robust training infrastructure that would provide further opportunities for pharmacists to develop their skills and promote lifelong learning are therefore required in the country. Enhanced pharmaceutical care services such as medication chart review, medication error reporting, antibiotic stewardship programmes, and pharmacovigilance activities are important clinical services that impact patient outcomes [2,37]. Prioritizing pharmacists’ involvement in these key services in Nigeria is crucial in order to safeguard patients’ health, ensure medication safety, and limit drug interactions.

To the best of our knowledge, this is the first study to provide a comparative overview of the available clinical pharmacy services within the various levels of care and across the six geopolitical regions of Nigeria. The national scope of the study is a key strength, given the paucity of data in the subject area. However, given that this was a non-random survey, our study estimates though accurate, are unlikely to be a precise representation of the population of interest. Also, this study relied on respondents’ report of the services available at their respective hospital. This approach is likely to have introduced bias associated with the use of self-administered questionnaires [38]. Despite these limitations, the broad similarities in service provision within the respective levels of care in our sample, provide an indication of the existing trends with respect to clinical pharmacy service availability across the states and geopolitical regions in the country.

## Conclusions

This study provides evidence that was previously lacking with respect to the availability of clinical pharmacy services in public sector hospitals in Nigeria. The disparity in the availability of these services across the various levels of care highlight the need to scale up the uptake of these services nationally. Our findings also emphasize the need to prioritize the integration of clinical pharmacy services within the primary healthcare system, to ensure equitable access to medicines expertise and contribute to universal health coverage. The evidence in this study can inform national policy planning and development in the pharmaceutical sector in Nigeria, particularly in relation to ensuring the availability of enhanced patient-oriented services.

## Data Availability

The authors confirm that the data supporting the findings of this study are available within the article

## Author contribution

- AU – Conceptualization, methodology, data curation, formal analysis, original draft preparation
- EN, EK, UO, MA, UII - Conceptualization, methodology, data curation, validation, writing review and editing
- BKL, KSL, KA, AM, OT - Methodology, data curation, validation, writing review and editing

## Acknowledgment

The authors would like to thank the Association of Hospital and Administrative Pharmacists of Nigeria (AHAPN), the Pharmaceutical Society of Nigeria (PSN) and Olutope Arinola Akinnibosun for assisting with the data collection in this study.

---

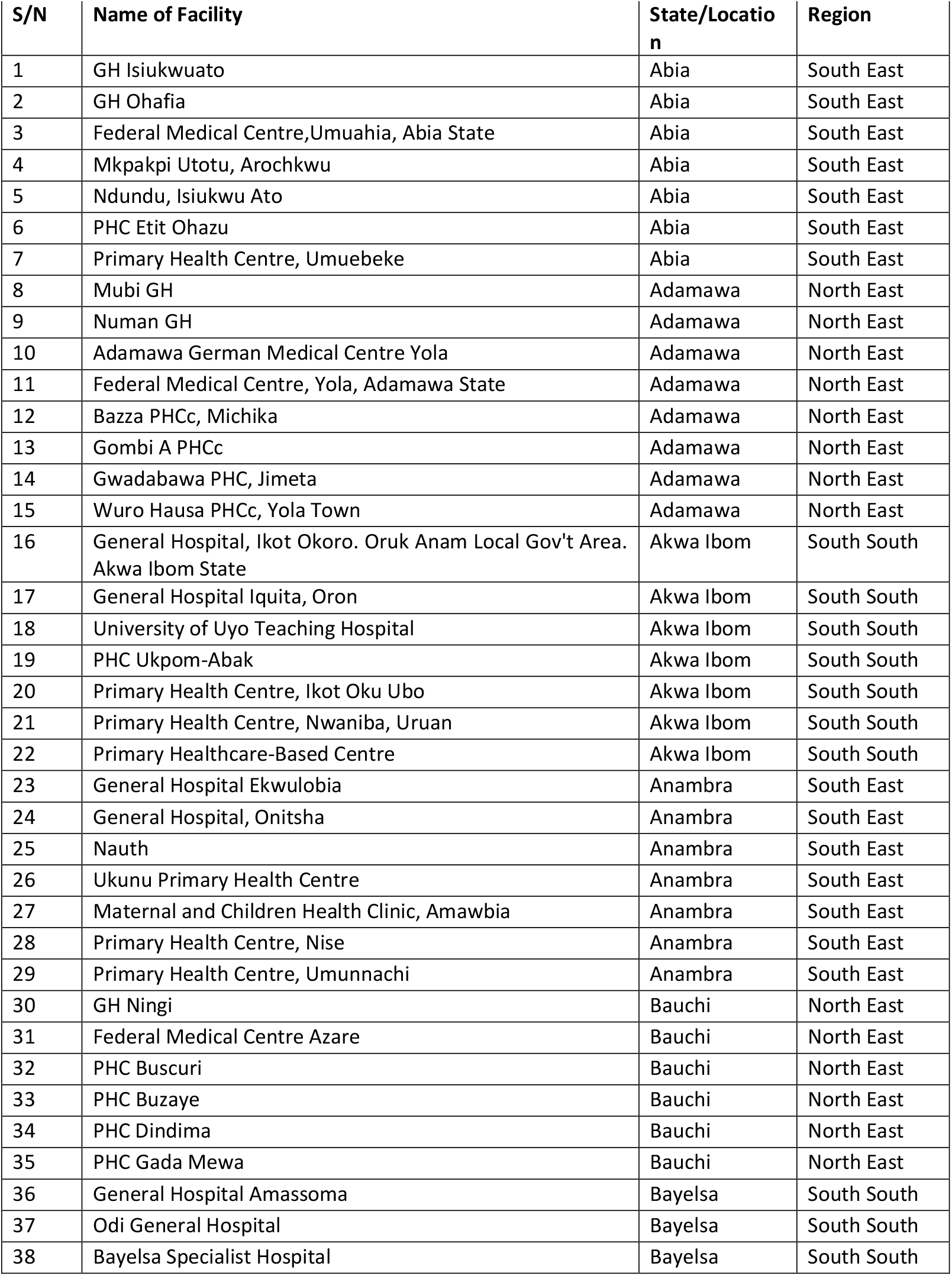
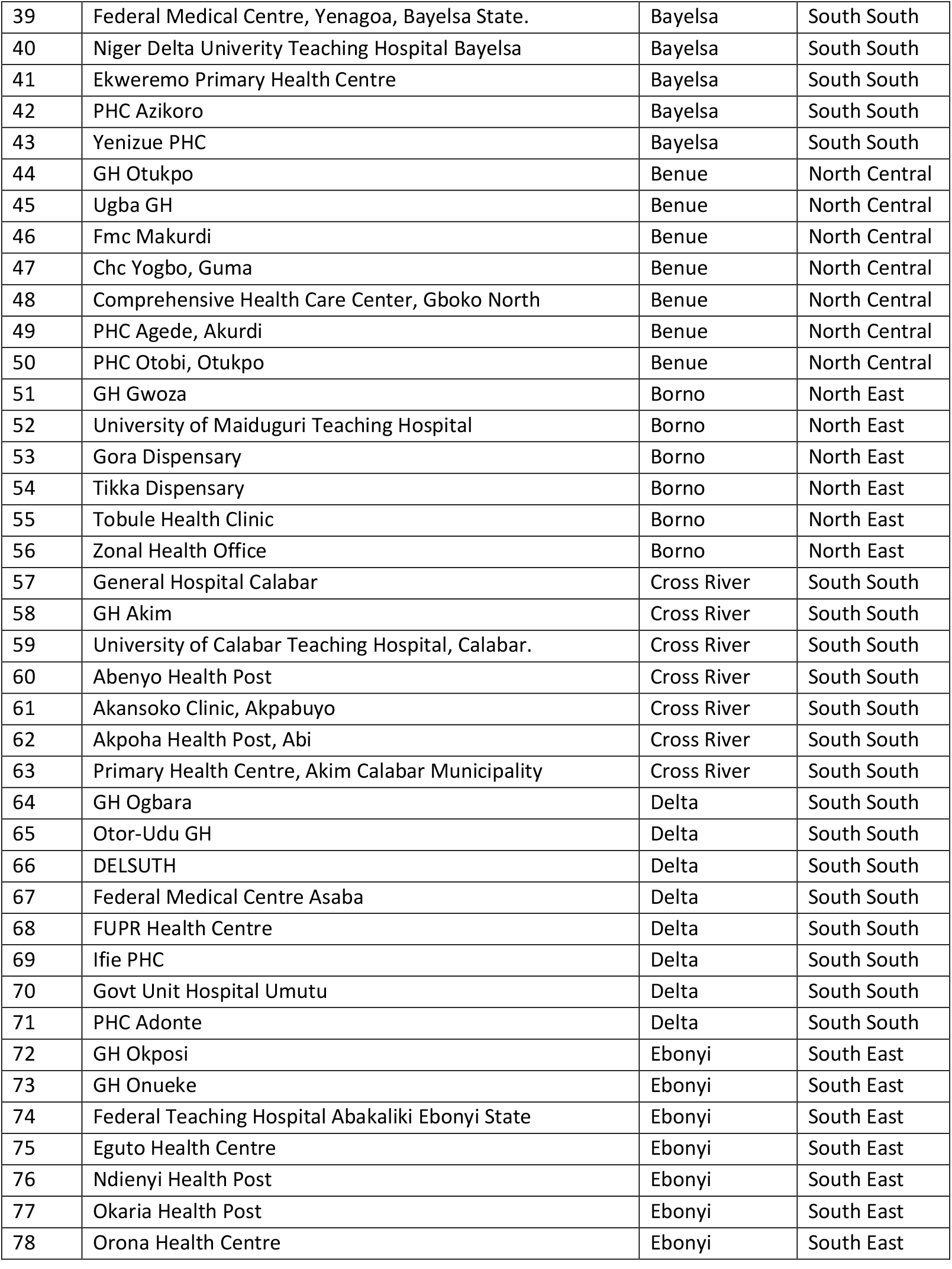
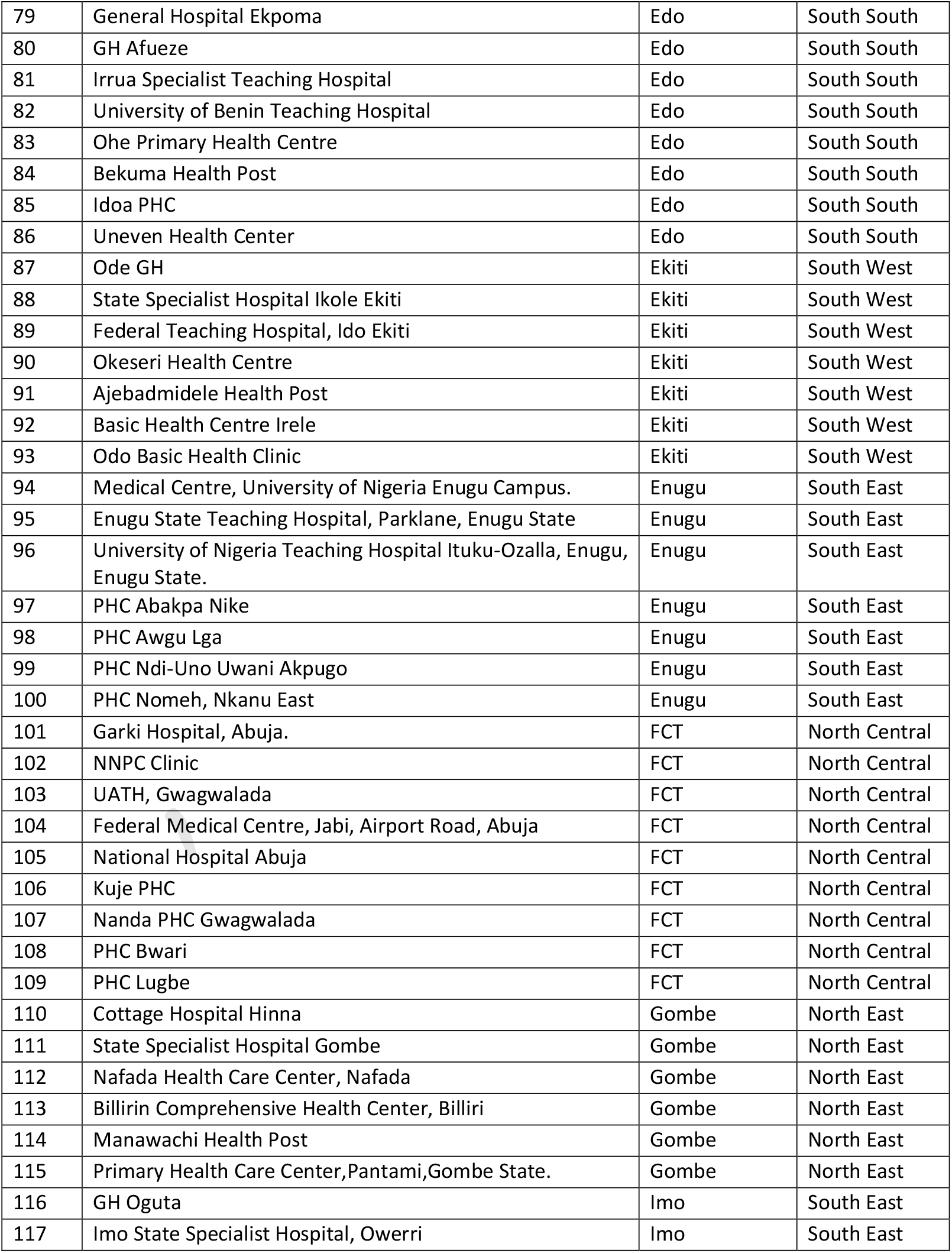
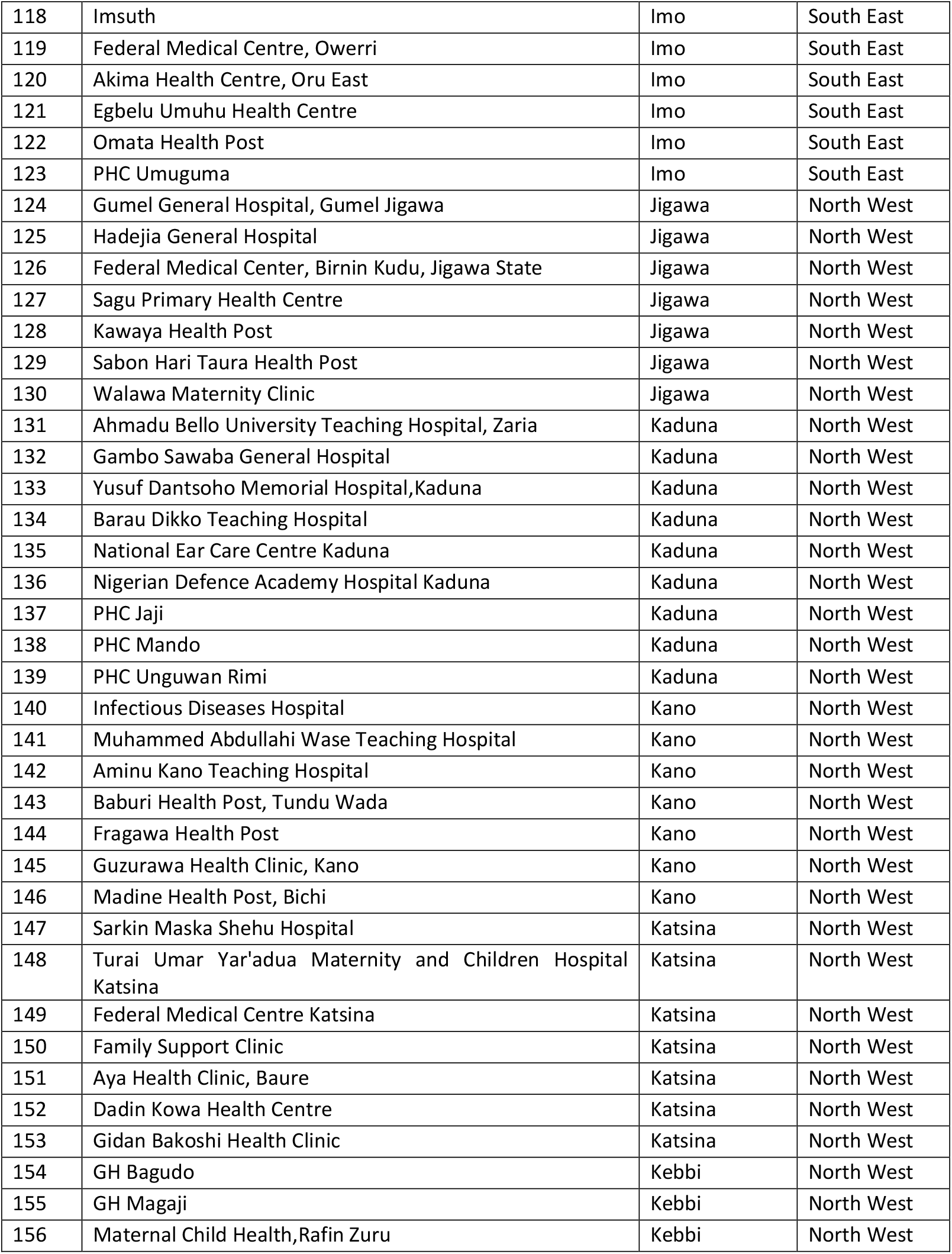
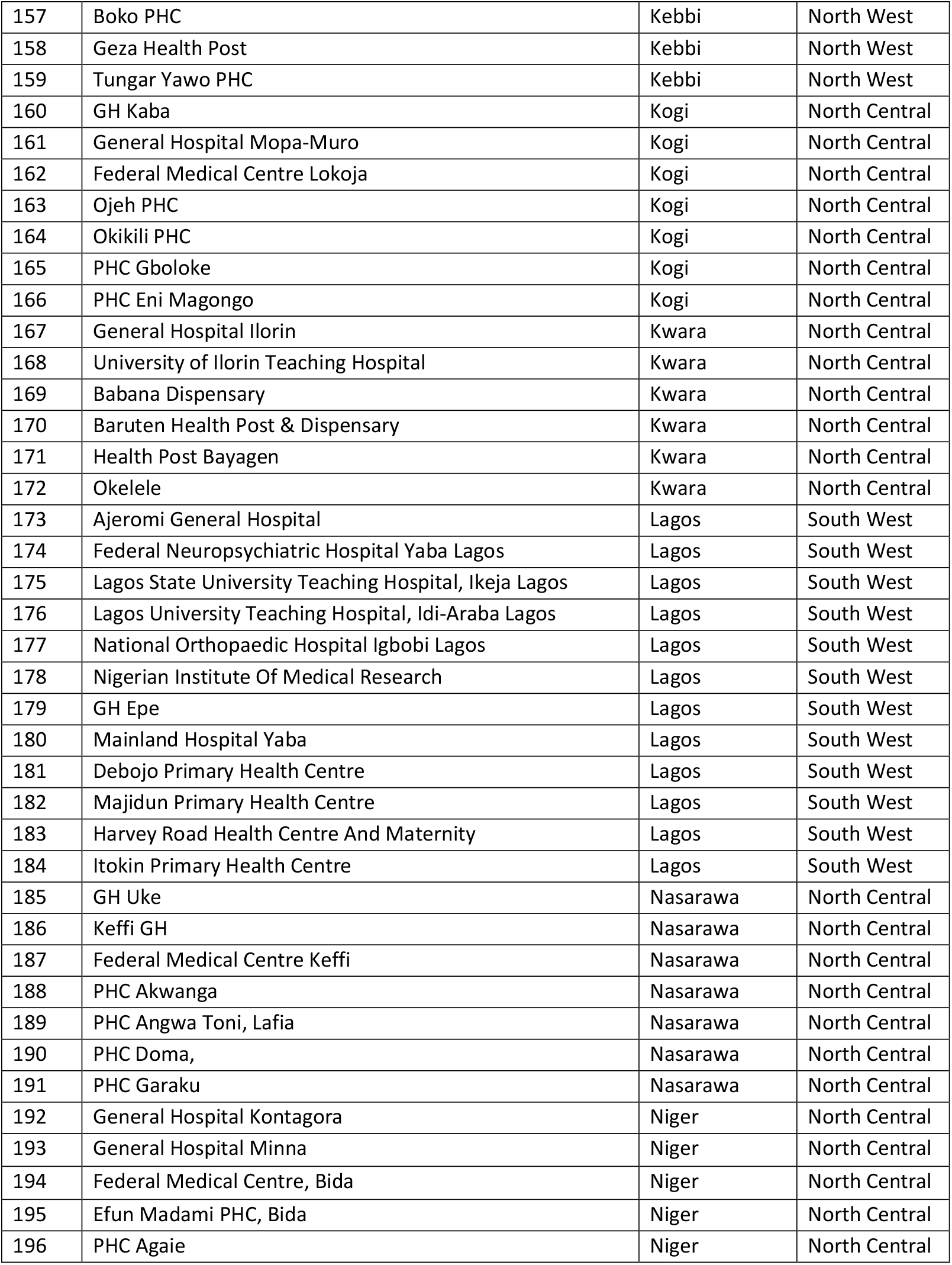
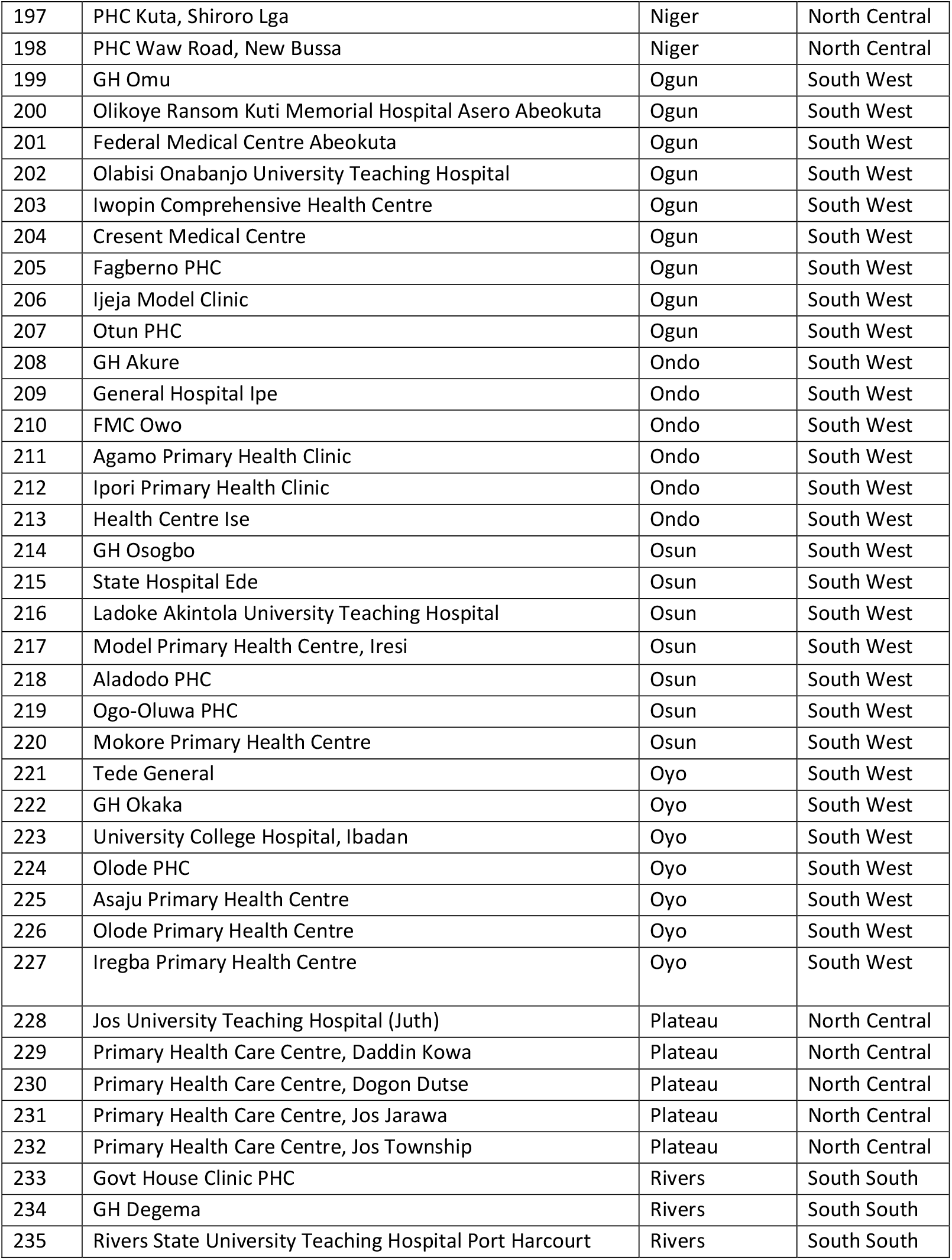
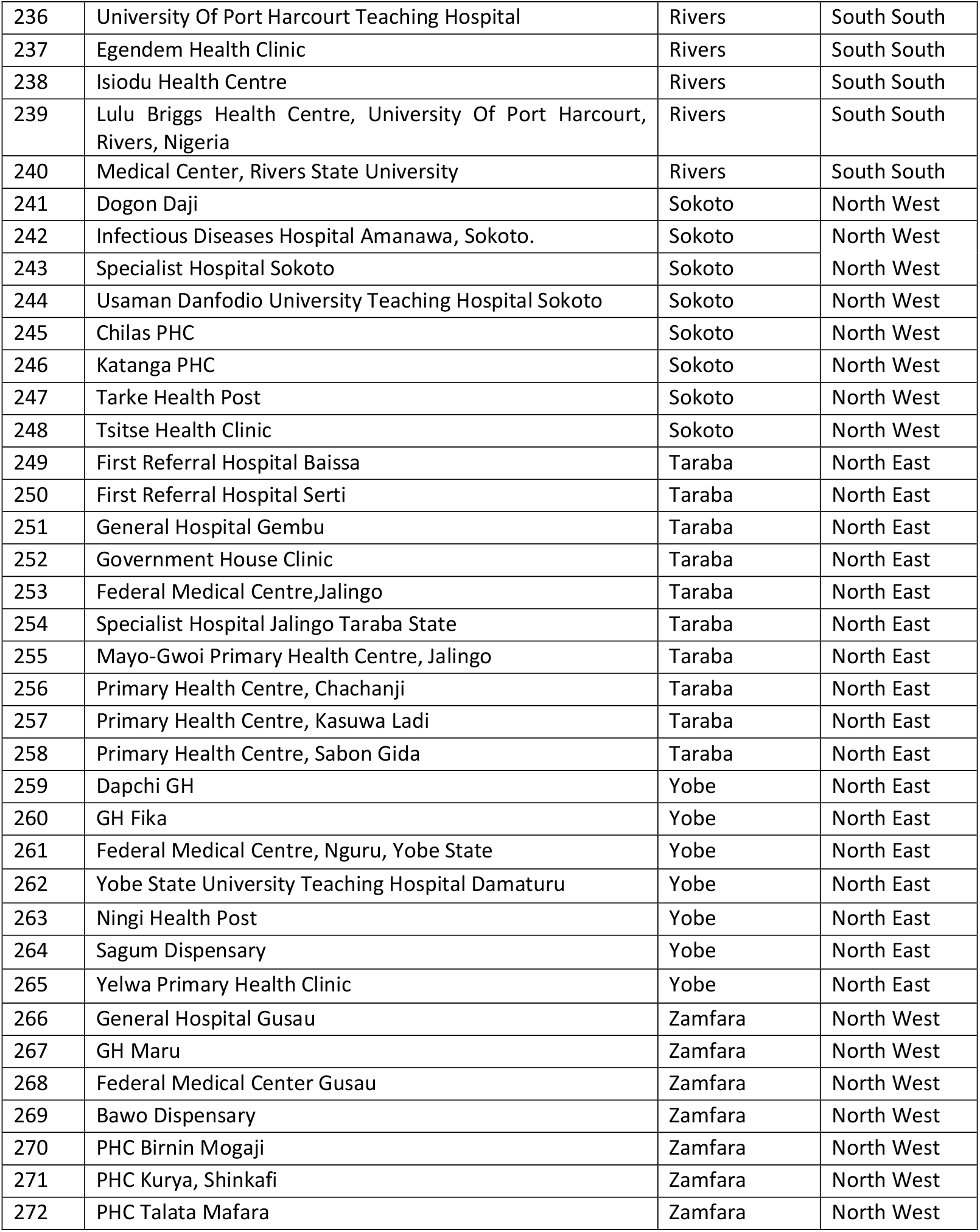

## References

1. Obayendo T. Presidency Approves Consultant Pharmacist Cadre in Public Service. Pharmanewsonline 2020.

2. Rotta I, Salgado TM, Silva ML et al. Effectiveness of clinical pharmacy services: an overview of systematic reviews (2000-2010). Int J Clin Pharm 2015;37:687–97.

3. Carter BL. Evolution of Clinical Pharmacy in the USA and Future Directions for Patient Care. Drugs Aging 2016;33:169–77.

4. Canadian Society of Hospital Pharmacists. Hospital Pharmacy in Canada Report 2016/17. Ottawa, Canada: Canadian Society of Hospital Pharmacists, 2018.

5. Proper JS, Wong A, Plath AE et al. Impact of clinical pharmacists in the emergency department of an Australian public hospital: A before and after study. Emerg Med Australas 2015;27:232–8.

6. Studer H, Boeni F, Messerli M et al. Clinical Pharmacy Activities in Swiss Hospitals: How Have They Evolved from 2013 to 2017? Pharm Basel Switz 2020;8, DOI: 10.3390/pharmacy8010019.

7. The Pharmaceutical Society of Ireland. Baseline Study of Hospital Pharmacy in Ireland: Final Report. Dublin, Ireland: The Pharmaceutical Society of Ireland, 2012:205.

8. Elliott RA, Perera D, O’Leary K. National survey of clinical pharmacy services and pharmacy technician roles for subacute aged-care inpatients. J Pharm Pract Res 2012;42:125–8.

9. Hambisa S, Abie A, Nureye D et al. Attitudes, Opportunities, and Challenges for Clinical Pharmacy Services in Mizan-Tepi University Teaching Hospital, Southwest Ethiopia: Health Care Providers’ Perspective. Adv Pharmacol Pharm Sci 2020;2020:e5415290.

10. Pawlowska I, Pawlowski L, Kocic I et al. Clinical and conventional pharmacy services in Polish hospitals: a national survey. Int J Clin Pharm 2016;38:271–9.

11. Yao D, Xi X, Huang Y et al. A national survey of clinical pharmacy services in county hospitals in China. PLOS ONE 2017;12:e0188354.

12. Auta A, Strickland-Hodge B, Maz J. Challenges to clinical pharmacy practice in Nigerian hospitals: a qualitative exploration of stakeholders’ views. J Eval Clin Pract 2016;22:699–706.

13. Bronkhorst E, Gous AGS, Schellack N. Practice Guidelines for Clinical Pharmacists in Middle to Low Income Countries. Front Pharmacol 2020;11, DOI: 10.3389/fphar.2020.00978.

14. Njuguna B, Berhane H, Ndemo FA et al. Scaling up clinical pharmacy practice in Africa: Current challenges and the future. JACCP J Am Coll Clin Pharm 2020;3:966–72.

15. International Pharmaceutical Federation. Revised FIP Basel Statements on the Future of Hospital Pharmacy. The Hague, Netherlands: International Pharmaceutical Federation, 2014:6.

16. Penm J, Moles R. Global developments in hospital pharmacy: the revised Basel statements. J Pharm Pract Res 2016;46:301–3.

17. Ekpenyong A, Udoh A, Kpokiri E et al. An analysis of pharmacy workforce capacity in Nigeria. J Pharm Policy Pract 2018;11:20.

18. Ma’aji HU, Khan F, Shuaibu A et al. Assessment of Hospital Pharmacy Services in North Western Nigeria. Niger J Pharm Sci 2018;17:90–7.

19. The World Bank. Population, total - Nigeria Data. 2019.

20. International Organization for Migration. Needs Assessment of the Nigerian Health Sector. Abuja, Nigeria: International Organization for Migration, 2014:47.

21. Kombe G, Fleisher L, Kariisa E et al. Nigeria Health System Assessment. Bethesda, MD: United States Agency for International Development and Abt Associates Inc., 2009.

22. National Primary Health Care Development Agency. Report of the Expert Group on Revitalization of Primary Health Care In Nigeria. Abuja, Nigeria: National Primary Health Care Development Agency, 2015:97.

23. Alliance for Health Policy and Systems Research. Primary Care Systems Profiles and Performance (PRIMASYS). Geneva, Switzerland: Alliance for Health Policy and Research, The Bill & Melinda Gates Foundation and World Health Organization, 2016:8.

24. Asuzu M. The necessity for a health system reform in Nigeria. J Community Med Prim Health Care 2004;16:1–3.

25. Federal Ministry of Health. Distribution of Hospitals and Clinics in Nigeria. Niger Health Facil Regist HFR 2017.

26. Olson CL. Comparative Robustness of Six Tests in Multivariate Analysis of Variance. J Am Stat Assoc 1974;69:894–908.

27. Eysenbach G. Improving the Quality of Web Surveys: The Checklist for Reporting Results of Internet E-Surveys (CHERRIES). J Med Internet Res 2004;6:e34.

28. Udoh A, Ernawati DK, Akpan M et al. Pharmacies and primary care: a global development framework. Bull World Health Organ 2020;98:809–11.

29. Alenoghena I, Aigbiremolen A, Abejegah C et al. Primary Health Care in Nigeria: Strategies and Constraints in Implementation. Int J Community Res 2014;3:74–9.

30. Christian Aid. Assessment of Primary Health Centres in Selected States of Nigeria: Summary Report of Findings from Christian Aid Supported Communities in Anambra, Benue, Kaduna, Plateau States and the Federal Capital Territory (FCT). Abuja, Nigeria: Christian Aid, 2015:38.

31. Tan ECK, Stewart K, Elliott RA et al. Pharmacist services provided in general practice clinics: a systematic review and meta-analysis. Res Soc Adm Pharm RSAP 2014;10:608–22.

32. Avery AJ, Rodgers S, Cantrill JA et al. A pharmacist-led information technology intervention for medication errors (PINCER): a multicentre, cluster randomised, controlled trial and cost-effectiveness analysis. Lancet Lond Engl 2012;379:1310–9.

33. Adibe M, Victoria U. Assessment of Hospital Pharmacy Services in South-Eastern Nigeria. Int J Pharmagenesis 2010;1:209–15.

34. Bilal AI, Tilahun Z, Gebretekle GB et al. Current status, challenges and the way forward for clinical pharmacy service in Ethiopian public hospitals. BMC Health Serv Res 2017;17:359.

35. Salim AA, Elhada AA, Elgizoli B. Exploring clinical pharmacists’ perception of their impact on healthcare in Khartoum State, Sudan. J Res Pharm Pract 2016;5:272.

36. Institute of Medicine. To Err Is Human: Building a Safer Health System. Washington DC: National Academy Press, 2000.

37. Ourghanlian C, Lapidus N, Antignac M et al. Pharmacists’ role in antimicrobial stewardship and relationship with antibiotic consumption in hospitals: An observational multicentre study. J Glob Antimicrob Resist 2020;20:131–4.

38. Demetriou C, Ozer BU, Essau CA. Self-Report Questionnaires. The Encyclopedia of Clinical Psychology. American Cancer Society, 2015, 1–6.

